# Operationalisation of the African Medicines Agency: Retrospective evaluation of the continental centralized pilot procedure – timelines to recommendation and national registration decisions

**DOI:** 10.64898/2026.04.22.26351547

**Authors:** Alex Juma Ismail, Lerato Moeti, Delese Mimi Darko, Stuart Walker, Sam Salek

## Abstract

**Background:** Regulatory inconsistency across African countries contributes to duplicative scientific assessments, prolonged approval timelines, and delayed access to essential medical products. To inform the operationalisation of the African Medicines Agency (AMA), the African Medicines Regulatory Harmonisation (AMRH) programme implemented Africa’s first continental pilot study for the scientific evaluation and listing of human medicinal products. This study evaluates the pilot’s procedural performance and examines how continental scientific opinions were translated into national regulatory decisions through reliance mechanisms.

**Methods and Findings:** A mixed-methods programme evaluation was conducted using regulatory datasets generated during the pilot study. Quantitative data included assessment timelines, GMP inspection outcomes and national post-listing regulatory actions. Qualitative thematic analysis was applied to governance documents and National Regulatory Authority (NRA) feedback to identify legal, institutional and procedural determinants influencing uptake. Of 64 expressions of interest, 24 products progressed to full evaluation and 12 received positive continental scientific opinions. Ten met the predefined performance target of ≤210 working days. Twenty-four GMP inspections identified no critical deficiencies and aligned with global regulatory benchmarks. National uptake demonstrated active reliance: full reliance (continental opinion as primary basis for national approval) for 7 products (58%); sequential reliance (continental assessment supplemented with targeted national queries) for 3 products (25%); and supplemented national review (separate national assessment undertaken) for 2 products (17%). Products with broader market strategies achieved registration in up to 23 African countries within a median of 77 working days post-listing. Variability in uptake reflected national legal authority, administrative requirements, and applicant submission strategies

**Conclusions:** The pilot study demonstrates the feasibility of a continent-wide regulatory assessment mechanism capable of producing trusted scientific outputs and enabling reliance-based national decision-making in Africa. While reliance was widely applied, heterogeneity in national procedures and administrative sequencing affected time to national registration. Findings provide empirical evidence to inform the AMA scale-up, highlighting the need for harmonised reliance pathways, streamlined administrative processes, and coordinated digital regulatory infrastructure.

**Author Summary:** **Why Was This Study Done?**

Access to essential, safe, and effective medicines in Africa is often delayed because regulatory systems across countries work independently and have different capacities. To strengthen regulatory efficiency, African institutions have established the African Medicines Agency (AMA) which will carry out continent-wide scientific assessments that countries can rely on when making their own regulatory decisions. However, since no evidence previously existed to show whether such a continental process could work in practice, there was a need to carry out such a study.

**What Did the Researchers Do and Find?**

Africa’s first pilot study of a continent-wide regulatory assessment process for human medicines was evaluated. Of 24 eligible applications, 12 products were listed following a positive scientific opinion issued after scientific assessment and manufacturing quality evaluation. Sixteen national regulatory authorities used the continental assessments directly when granting marketing authorisations, reducing duplication and accelerating decision-making. Countries varied in how quickly and extensively they adopted the continental recommendations, depending on national laws, administrative procedures and capacity.

**What Do These Findings Mean?**

A continental assessment system is feasible in Africa that can provide the basis for reliance-based national decisions. To make this approach fully effective under the African Medicines Agency, countries will need clearer reliance legislations and pathways, streamlined administrative processes, and systems that connect continental, regional and national regulatory procedures.

## 1. Introduction

Access to safe, effective, and quality-assured medical products remains inconsistent across Africa where regulatory systems are fragmented and vary widely in capacity. Nine out of 54 National Regulatory Authorities (NRAs) in the region operate at a maturity level sufficient to perform core regulatory functions consistently according to the WHO Global Benchmarking Tool [1–2]. Such variability contributes to the duplication of scientific assessments, prolonged and unpredictable registration timelines and delays in access to essential medicines problems well documented across low- and middle-income countries [3–5].

Globally, regulatory reliance has emerged as a pragmatic strategy to reduce duplication while maintaining national sovereignty. The WHO defines reliance as the use of another authority’s scientific assessment to inform national decision-making supported by good governance and trust between regulators [6]. Evidence shows that structured reliance can improve efficiency, strengthen regulatory convergence and optimise limited resources in constrained settings [7]. International collaborative models including the European Medicines Agency’s coordinated assessment network, WHO Prequalification, and the WHO Collaborative Registration Procedure illustrate how shared scientific opinions can accelerate access when uptake mechanisms are robust [8–11].

In Africa, reliance efforts have been pursued mainly through regional harmonisation initiatives such as the Southern African Development Community’s ZaZiBoNa process, the East African Community’s joint assessment procedure, and the West African Health Organization’s ECOWAS mechanisms [12–15]. These initiatives demonstrate that work-sharing is feasible and can shorten timelines; however, evaluations highlight persistent challenges including voluntary participation, inconsistent national uptake, limited legal provisions for reliance, and structural capacity constraints [5,16].

The creation of the African Medicines Agency (AMA) represents a major shift toward continent-wide coordination. The AMA Treaty and corresponding African Union decisions mandate AMA to support joint scientific assessments, coordinated inspections, and structured reliance pathways while respecting national regulatory authority [17–20]. This institutional architecture builds on the African Medicines Regulatory Harmonisation (AMRH) programme and continental policy frameworks that call for reducing duplication and ensuring equitable access across countries with differing regulatory capacities [21].

To generate empirical evidence for AMA operationalisation, the AMRH programme implemented the first continental pilot study for the scientific evaluation and listing of human medicinal products between 2023 and 2025. The pilot study tested whether harmonised, continent-wide assessment and Good Manufacturing Practice (GMP) inspection processes could produce credible scientific opinions and whether NRAs would translate these into national regulatory decisions. Although regional initiatives have produced shared assessments, no prior study has analysed a continental procedure or systematically mapped its downstream national uptake.

The aims of the study were to evaluate the continental listing pilot study as a regulatory systems intervention to test: (1) the feasibility and performance of the continental assessment and inspection procedure; (2) how NRAs apply reliance to convert the issued continental scientific opinions into national regulatory decisions or product registration; and (3) the legal, procedural and institutional factors influencing uptake. These findings provide the first empirical evidence on continental-level assessment and reliance in Africa and offer critical insights for AMA’s centralized process design and future implementation. Furthermore, this study was designed to generate evidence to support institutionalisation and sustainability of reliance-based regulatory functions to be implemented under the AMA.

## 2. Methods

### 2.1 Study Design

A mixed-methods programme evaluation of Africa’s first continental centralized procedure for the scientific evaluation and listing of human medicinal products was conducted and implemented under the AMRH initiative. The pilot study was conceived as a regulatory systems intervention designed to test core functions anticipated for the AMA including joint scientific assessment, coordinated Good Manufacturing Practice (GMP) inspections, and reliance-based national decision-making. Mixed-methods designs are appropriate for complex regulatory and health-system interventions where quantitative performance metrics must be interpreted alongside institutional and contextual factors influencing implementation [26,28].

#### Setting: The Continental Listing Centralized Procedure

The pilot study was operated through AMRH governance structures. Scientific assessments and Good Manufacturing Practice (GMP) inspections of manufacturing sites involved were conducted by experts from African National Regulatory Authorities (NRAs) under the oversight of the Evaluation of Medicinal Products Technical Committee (EMP-TC) and the GMP Technical Committee (GMP-TC) respectively. The final decisions were issued by the AMRH Steering Committee which led to products being issued with a positive opinion being listed in the Green Book, a process referred to as *Continental Listing*. Guidelines, Procedures, templates, eligibility criteria and performance benchmarks were predefined using the WHO Good Reliance Practices and African Union regulatory policy frameworks [6,19]. The continental listing was to provide a consolidated scientific opinion but not to grant marketing authorisation in participating countries for the national NRAs retain full legal authority.

### 2.2 Data Sources

Three categories of routinely generated regulatory data were analysed:

1. Continental procedural data: including expressions of interest, dossier screening and validation outcomes, scientific assessment reports, GMP inspection reports, committee deliberations, and listing decisions maintained by the AMRH Secretariat [22].
2. National regulatory data: country-reported actions following continental scientific opinion, including reliance modality, national submission status and marketing authorisation decisions [22].
3. Regulatory policy and procedural documents: including the AMA Treaty, continental guidance, and reliance frameworks from WHO and the International Coalition of Medicines Regulatory Authorities (ICMRA) [6,10,19].

Data were accessed for research purposes between 20 January 2026 and 26 March 2026. The authors did not have access to any information that could identify individual patients or human participants during or after data collection, as the dataset consisted solely of de-identified regulatory procedural data, product-level assessment outcomes, and aggregate national uptake statistics.

### 2.3 Quantitative Analysis

Descriptive statistics were used to characterise the pilot study pipeline, assessment timelines, GMP inspection outputs and post-listing decision national uptake. Official AMRH procedural timelines were measured from dossier acceptance to Steering Committee decision, consistent with pilot study protocol definitions. Total elapsed time from dossier submission to listing was estimated separately. GMP inspection findings were categorised by deficiency type and severity following harmonised international definitions.

National uptake was analysed at the product–country level. Reliance modalities were categorised as:

- Full reliance: the continental scientific opinion was the primary basis for national approval, with only administrative checks added.
- Supplemented reliance: continental assessment was implemented with the addition of specific national queries or additional procedural steps.
- Sequential review: a separate national scientific assessment was undertaken following the review by the AMRH

Missing national data were resolved through cross-checking with NRAs and the AMRH Secretariat’s Evaluations coordinator where feasible.

### 2.4 Qualitative Analysis

To understand the legal, institutional, and procedural factors influencing national uptake of continental scientific opinions, a retrospective qualitative document analysis of regulatory records generated during and immediately following the pilot study was conducted. Data sources included: (i) written feedback from National Regulatory Authorities (NRAs) submitted to the AMRH Secretariat following continental listing (ii) governance records from EMP-TC, GMP-TC and Steering Committee meetings (iii) national reliance policies or procedural guidelines referenced by NRAs during post-listing communications; and (iv) correspondence between applicants and NRAs where relevant to understanding regulatory decisions.

The analysis followed the six-phase framework for thematic analysis described by Braun and Clarke [30]. This approach was selected for its flexibility and suitability for identifying patterns across diverse textual data sources. The phases were operationalised as follows:

- **Phase 1. Familiarisation with data:** All documents were reviewed iteratively by the lead author to gain an understanding of the content and to note initial observations.
- **Phase 2. Generating initial codes:** A deductive coding structure was developed based on realist evaluation concepts context, mechanism, outcome [23,24] and implementation science frameworks describing determinants of regulatory uptake [25]. Codes captured references to legal authority for reliance, procedural alignment, administrative requirements, resource constraints, and institutional behaviours. Inductive coding allowed additional themes to emerge from the data.
- **Phase 3. Searching for themes:** Codes were grouped into candidate themes through iterative discussion among the research team. Themes were organised around factors that appeared to enable or impede uptake.
- **Phase 4. Reviewing themes:** Themes were checked against the original data and refined to ensure internal coherence and external distinctiveness. Discrepancies were resolved through team discussion.
- **Phase 5. Defining and phrasing of themes:** Final themes were clearly defined, with each linked to illustrative data excerpts. Themes focused on legal authority, procedural alignment, administrative sequencing, and institutional factors influencing reliance.
- **Phase 6. Producing the report:** Findings were integrated into the interpretation of quantitative uptake patterns and are presented in the discussion section.

Coding was managed using a combination of spreadsheet software and manual annotation. To enhance interpretive validity, findings were triangulated across data sources and reviewed by co-authors with direct involvement in the pilot study [29]. No qualitative analysis software was used; coding was conducted manually by the lead author with regular team validation.

### 2.5 Ethical Considerations

Ethical approval for this study was granted by the University of Hertfordshire’s Health, Science, Engineering and Technology Ethics Committee with UH protocol number is: 2070 ST HSET 2026. The study used secondary institutional data generated through routine regulatory operations. The study was also consistent with WHO guidance for programme evaluation [26]

## 3. Results

### 3.1 Data Characteristics of the Continental Listing Pilot Study

The pilot study received 64 expressions of interest (EOIs) from applicants. Following screening against predefined eligibility criteria, 24 products (38%) were shortlisted for full continental evaluation. The reduction from 64 to 24 was due to some EOIs not meeting product eligibility requirements (e.g., product type, inability to provide electronic Common Technical Documentation (e-CTD) formats); others lacked signed commitment to submit dossier within the pilot study timeline; and some applicants were unable to commit to the required GMP inspection schedule including readiness for onsite inspections at their respective manufacturing sites.

Of the 24 products that entered full evaluation, 21 proceeded to all cycles of scientific assessment, while three were withdrawn prior to completion of the first assessment cycle. These withdrawals occurred because applicants could not meet the requirement to host an onsite GMP inspection within the pilot timeframe due to operational challenges of coordinating inspections across multiple jurisdictions and manufacturing sites.

By the conclusion of the pilot, 12 products received a positive continental scientific opinion and were included in the continental list known as Green Book, 2 products received a negative opinion and were rejected. The remaining seven fully evaluated products did not progress to listing due to: unresolved scientific deficiencies identified during the three-cycle assessment process; negative GMP inspection outcomes requiring corrective actions that could not be completed within the pilot period or voluntary withdrawal by applicants following scientific feedback from the technical committees.

This pipeline was rigorous, multi-stage nature designed into the continental assessment process where only those products meeting all scientific quality, safety and efficacy requirements at each step proceeded to final listing.

### 3.2 Scientific Assessment

#### 3.2.1 Consolidated Scientific Assessment

All products entering evaluation underwent a harmonised three-cycle scientific assessment overseen by the EMP-TC. Each application receiving a positive outcome was issued a single consolidated scientific opinion covering quality (including GMP compliance), safety, and efficacy.

#### 3.2.2 Timelines for Continental Assessment and Listing

The AMRH procedural workflow demonstrated consistent performance for most products achieving continental listing. The official AMRH timeline, measured from dossier acceptance to publication on the Green Book ranged from 148 to 233 working days. (Median value of 165 days) Ten of the 12 products (83%) met the ≤210 working-day target (**Table 1**).

**Table 1.** Summary of Timelines from Dossier Acceptance to Continental Decision by Product (n=12)

| **Product Name** | **Therapeutic Class** | **AMRH Time to Recommendation (Working Days)** |
| --- | --- | --- |
| 1. EasySix Vaccine | Vaccine (DTwP-HepB-Hib-IPV) | 148 |
| 2. Bavencio | Monoclonal Antibody (Oncology) | 149 |
| 3. Keytruda | Monoclonal Antibody (Oncology) | 154 |
| 4. Gardasil 9 (Syringe) | Vaccine (HPV) | 153 |
| 5. Gardasil 9 (Vial) | Vaccine (HPV) | 153 |
| 6. Zincol | Mineral Supplement (Zinc) | 155 |
| 7. Truqap 160 mg | Small Molecule (Oncology) | 176 |
| 8. Truqap 200 mg | Small Molecule (Oncology) | 176 |
| 9. Strensiq 40 mg/ml | Enzyme Replacement (Metabolic) | 193 |
| 10.Strensiq 100 mg/ml | Enzyme Replacement<br>(Metabolic) | 193 |
| 11. Soliris | Monoclonal Antibody<br>(Haematology/Immunology) | 223 |
| 12. IMFINZI | Monoclonal Antibody<br>(Oncology) | 233 |
| **All Products (n=12)** |  |  |
| **Median** |  | **165** |
| **Range** |  | **148 – 233** |

The continental pilot study successfully established a functional assessment pathway and generated comprehensive timeline data for the 12 products that received a positive scientific opinion and were listed in the AMRH Green Book.

A stage-wise analysis of the pilot’s performance against its planned benchmarks (**Table 2**) indicates end-to-end continental scientific recommendation achieved a median of 198 working days within the ≤210-day target though the observed range extended to 242 working days.

**Table 2.** *Summary of Key Timelines for the Continental Listing Pilot Study.

| Indicator | Planned Benchmark<br>(Calendar Days) | Observed Median<br>(Working Days) | Observed Median<br>(Calendar Days) | Observed Range<br>(Working Days) | Observed Range<br>(Calendar Days) |
| --- | --- | --- | --- | --- | --- |
| Technical screening | $\leq 15$ | 2 | ~3 | 1 – 3 | ~1 – 4 |
| Scientific assessment<br>(EMP-TC) | $\leq 150$ | 138 | ~193 | 112 – 176 | ~157 – 246 |
| GMP inspection cycle | $\leq 60$ | 55 | ~77 | 42 – 91 | ~59 – 127 |
| End-to-end listing timeline | ≤210 | 198 | ~277 | 171 – 242 | ~239 – 339 |

\*Note: Calendar days are approximate conversions based on a standard 5-day working week (1 working day = 1.4 calendar days). Actual calendar days may vary depending on public holidays and weekends.

#### 3.2.3 GMP Inspection Outcomes

A total of 24 GMP inspections were conducted for manufacturing sites linked to the evaluated products. These included onsite, remote, and desk-based inspections, selected according to risk and site accessibility.

Of these: 23 inspections were successfully completed, one inspection of the drug substance was not completed following a withdrawal by the applicant from the list of its suppliers during the last cycle of evaluation of the product, no critical deficiencies were identified, 93% of all deficiencies were classified as *major*, and common deficiency themes included contamination control, data integrity, and quality system implementation. Inspection findings from continental inspections aligned with trends observed by major international regulators.

### 3.3 National Uptake of Scientific Recommendations

National uptake was evaluated at the product–country level for all 12 listed products. Following continental listing, national regulatory authorities (NRAs) demonstrated varying degrees of reliance on the continental decision: seven products (58%) were processed through full reliance resulting in immediate national registration, three products (25%) received supplemented reliance with additional national queries, and two products (17%) underwent sequential national review.

Under full reliance, the seven products Gardasil 9 (syringe and vial), Keytruda, Truqap (160 mg and 200 mg), Zincol and Bavencio achieved national registration in a total of 31 countries, with a median time to national decision of 77 working days (range: 11–116 days). Gardasil 9 alone accounted for 20 of these registrations, Keytruda was registered in 7 countries, while Truqap and Zincol each secured registration in one country following continental listing. Bavencio contributed two registrations (Tanzania and Uganda) under this pathway.

In contrast, products processed through supplemented reliance experienced substantially longer timelines. As of February 2026, 10 months after the March 2025 listings, Bavencio had been registered in only 2 of its 8 target countries with applications under evaluation in the remaining 5 countries and one country having issued a positive recommendation pending administrative finalisation. For products undergoing sequential national review Soliris and Strensiq no national registrations had been recorded with applications still under evaluation more than 10 months post-listing.

Uptake varied across products and across countries, reflecting differences in national procedures, administrative requirements, and implementation of reliance pathways. This analysis provided the basis for understanding why some products achieved rapid uptake through full reliance while others experienced delays due to supplemented reliance or sequential review, and for identifying the contextual factors that may shape future AMA operations.

### 3.4 National Uptake and Registration Outcomes Following Continental Listing

Following continental recommendation, there were 18 countries that registered Gardasil 9 (Vaccine – HPV) within 11-116 working days from the continental listing. However, there were three countries (Eritrea, Liberia and Uganda) for which the registration of Gardasil 9 was under evaluation. In addition, there were six countries (i.e. Cote d’ Ivoire, Gambon, Mauritius, South Africa, Tanzania and Ethiopia) that registered Keytruda for which information regarding duration from continental listing to NRA registration was not available. Furthermore, three countries (i.e. South Africa, Tanzania and Ethiopia) which pre-registered Keytruda for which time from continental listing and NRA registration was not relevant. All countries with pending applications as of February 2026 had exceeded the 90-day timeline, indicating systemic delays in those jurisdictions. In addition, only three countries (Ghana, Tanzania and Zambia) met the 90 working days target for national review, while six countries exceeded this timeline with decision taking up to 160 working days. This result is summarised in Table 3.

**Table 3.** Summary of Timelines from Continental Recommendation to National Decision by Country (n=23 countries)

| **item** | **Value** |
| --- | --- |
| Countries with Post-Listing Registrations (n) | 12 |
| Median Time to National Decision (Working Days) | 77 |
| Range (Working Days) | 11 – 116 |
| Countries with Registrations (Date Not Specified) | 8 |
| Countries Under Evaluation | 3 |
| Countries with Pre-Listing Registrations | 4 |

For the 12 countries with documented post-listing registration dates, the median time from continental listing to national decision was 77 working days (range: 11 - 116 days). Tanzania achieved the fastest registration at 11 working days while Namibia required 116 working days. An additional 8 countries granted registration but did not specify the decision date in their feedback to the Secretariat. Three countries (Eritrea, Liberia, Uganda) had applications under evaluation as of February 2026.

Besides Gardasil 9 and Keytruda, other products demonstrated varying uptake patterns. Truqap secured registration in South Africa following continental listing while Zincol achieved registration in Tanzania; one of its six target countries. Bavencio was registered in two countries (Tanzania and Uganda) under the full reliance pathway. For EasySix Vaccine, both target countries (Nigeria and Egypt) issued positive scientific recommendations based on the continental assessment, with final administrative steps pending. Products undergoing sequential national review Soliris and Strensiq remained under evaluation with no registrations recorded as of February 2026. Several NRAs, particularly in South Africa, explicitly referenced continental application identifiers in national submissions for products such as Bavencio, Keytruda, and Truqap, demonstrating operational integration of the continental system into national workflows.

## 4. Discussion

This evaluation provides the first empirical evidence on the feasibility and performance of a continent-wide scientific assessment and listing procedure for human medicinal products in Africa. The results demonstrate that consolidated continental scientific opinions can be produced efficiently, achieve broad national uptake and meaningfully support reliance-based decision-making across diverse regulatory systems. These findings offer practical insights for the operationalisation of the African Medicines Agency (AMA) whose mandate includes facilitating joint assessments, coordinated inspections and reliance-supported national decisions [17–20].

The three key principles that emerged from the study were, as follows: First, the continental scientific assessment process operated predictably and within defined performance expectations for ten of the twelve listed products, with 83% meeting the ≤210 working-day benchmark. This places the continental model within time ranges comparable to or faster than many national and regional procedures historically observed in Africa and across low- and middle-income countries [3–5]. The harmonised GMP inspection programme, drawing on risk-based and multimodal inspection modalities, generated findings consistent with international regulatory patterns [8] reinforcing the technical credibility of the assessments.

Second, national uptake of continental outputs was substantial. Most listed products were adopted through reliance pathways at the national level 58% through full reliance and 25% through supplemented reliance. This stands in contrast to earlier regional harmonisation efforts such as ZaZiBoNa, the East African Community joint assessment, and the ECOWAS mechanism, where national translation of joint assessments has been uneven or delayed [12–16]. The breadth and speed of uptake observed here suggest that centrally coordinated continental assessments may carry heightened legitimacy and utility for NRAs.

Third, end-to-end timelines from submission to national registration were heavily influenced not by the scientific assessment itself, but by administrative sequencing, including dossier intake intervals and the timing of Steering Committee meetings. A stage-wise analysis found that variability was greatest in the scientific assessment phase, largely due to assessment type and number of query cycles. However, the extended overall timelines observed were primarily attributable to procedural rather than scientific steps. Scientific assessments including dossier evaluation, GMP inspections, and technical committee deliberations were completed within or near their planned benchmarks (e.g., scientific assessment median of 138 working days against a 150-day target). The additional delays accumulated through applicant response times, fixed scheduling intervals for Steering Committee meetings, and administrative processing between dossier submission and formal acceptance.

These findings align with global evidence that reliance-based regulatory processes can improve efficiency, avoid duplication, and expand regulatory reach when supported by consistent procedures and transparency [6,7,10]. The continental approach shares characteristics with established models such as the European Medicines Agency’s coordinated network and WHO’s Prequalification and Collaborative Registration Procedure (CRP) both of which have demonstrated accelerated national regulatory uptake when assessments are jointly recognised [8–11].

A comparison with the EMA’s centralised procedure provides a useful benchmark for the AMRH pilot study. The EMA system, which has evolved over decades, represents a mature regulatory network where a single scientific assessment leads to simultaneous national decisions across all EU Member States [8,18].

Of the 12 products listed through the continental process, 9 (75%) have received EMA central approval. This includes all five monoclonal antibodies (Bavencio, Keytruda, Soliris, IMFINZI) both vaccines (Gardasil 9), both enzyme replacement products (Strensiq) and the small molecule oncology product Truqap. The two products without EMA approval EasySix, Zincol, and the withdrawn generics represent categories not typically submitted for EMA centralised assessment [32].

The timelines in working days observed in the AMRH pilot study (median 165 working days; range 148 - 233 days) are comparable to and in some cases faster than the EMA standard review timeline of 210 days [8]. The achieved 148 - 154 days review timeline aligns with EMA’s accelerated assessment pathway of 150 days, which is reserved for products of major public health interest [8]. This suggests that the African continental procedure when operating optimally, can achieve review speeds consistent with mature regulatory systems.

The WHO CRP provides another relevant comparator, as both mechanisms enable national regulatory authorities to rely on assessments conducted by trusted reference bodies [11]. As of 2025, 69 countries participate in the WHO CRP for medicines and nearly 1,200 product registrations have been facilitated through the procedure [9,11].

A recent case study illustrates the potential speed of the CRP pathway. Lenacapavir, a novel HIV prevention product, was approved in Zambia in 12 working days and in Zimbabwe in 18 working days. This rapid registration was as a result of reliance being placed on the EMA EU-M4all (formerly known as Article 58 – a regulatory procedure that allows the EMA to assess and provide scientific opinions on high-priority human medicines and vaccines intended for use outside the European Union) procedure and subsequent WHO prequalification [9,11]. These timelines demonstrate that when reliance is placed on Collaborative Registration procedures (CRP), then this can result in exceptionally rapid national approval, as was seen in the case of Zambia and Zimbabwe. These results also demonstrate that following the pilot study assessment, products can be rapidly registered in national regulatory authorities such as Tanzania (11 working days) and 30 working days in Zambia.

The WHO prequalification status provides an additional benchmark of global regulatory recognition. Since its establishment in 2001, the WHO Prequalification Programme has become a gold standard for assuring the quality, safety and efficacy of priority medicines, particularly those procured by United Nations agencies [9,18]. For example, Gardasil 9 and EasySix vaccine were prequalified by the WHO and subsequently reviewed rapidly in a national regulatory agency.

Regional harmonisation initiatives in Africa, including the East African Community (EAC) joint assessment procedure [13], the Southern African Development Community (SADC) collaborative medicines registration initiative [12], and the West African Health Organization (WAHO) harmonisation initiative [14], have made important progress in building regulatory capacity and facilitating work-sharing [15,16]. However, these subregional mechanisms typically achieve national uptake across a smaller geographic footprint, given their focus on specific economic communities [12,15]. The continent-wide reach of the AMRH pilot study with 27 countries targeted and 20 registrations achieved within 148 – 233 working days for leading products suggests that a pan-African mechanism may generate a degree of perceived harmonisation and authority beyond that achievable through subregional structures alone [16,17].

The fact that most listed products fall outside WHO’s traditional prequalification scope underscores the complementary nature of the two systems [9,18]. WHO PQ serves critical public health procurement needs for vaccines, HIV, TB, malaria, and reproductive health products [9]. The AMRH mechanism, in contrast, was meant to addresses the full spectrum of innovative medicines including oncology products, biologics, and rare disease treatments requiring market access across Africa [17,19,21]. This complementarity aligns with the African Union’s broader vision for the African Medicines Agency as a specialised agency capable of addressing the full range of regulatory needs across the continent [17,19,20].

These comparisons validate that the pilot study achieved technical equivalence with global regulatory standards while establishing a novel model for continental cooperation [17,22]. The finding that 75% of listed products already meet stringent EU regulatory requirements [8], and that key products also meet WHO prequalification standards [32,33], further reinforces the quality and credibility of the African assessment outcomes. As the African Medicines Agency moves toward full operationalisation, these results provide a strong foundation for the design and implementation of a sustained regulatory harmonisation and reliance across the continent [17,21,22].

The findings of this study have identified a number of implications for the operationalisation of the African Medicines Agency:

**First**, the strong uptake of continental assessments demonstrates that NRAs are willing to rely on shared scientific outputs when these are delivered through clear, transparent, and well-governed procedures. Embedding reliance pathways into national legislation and standard operating procedures may reduce variability in uptake and enhance predictability.

**Second**, administrative phases not scientific processes were the major contributors to extended end-to-end timelines. Streamlining dossier intake processes, standardising validation requirements, and ensuring more frequent or flexible scheduling of governance decisions could substantially reduce delays without compromising scientific quality.

**Third**, the pilot illustrates the value of coordinated GMP inspection capacity for a continent with diverse regulatory maturity levels [1,3]. AMA’s mandate to strengthen inspection cooperation [18–20] could leverage this approach to reduce duplication, support risk-based resource allocation, and align with global inspection norms.

**Fourth**, manufacturers demonstrated significant geographic breadth in their market-entry strategies, with several products targeting more than 20 African countries. A functional continental assessment mechanism may therefore encourage broader market participation and improve availability of quality-assured products across the AU.

Together, these implications point to a pragmatic pathway for AMA: institutionalise the procedural elements that worked, harmonise those that introduced variability, and reinforce the governance and digital systems required for efficient cross-country coordination [20,26].

### Strengths and Limitations

This study has several strengths. It draws on complete procedural and decision-making records generated by the AMRH Secretariat, enabling full visibility into assessment timelines, inspection outputs, and governance structures. It also incorporates verified post-listing national data, allowing direct observation of reliance patterns and registration outcomes across multiple national regulatory authorities.

The limitations include the pilot’s restricted number of products, which limits statistical generalisability. National uptake data reflects the period of observation: outcomes for products listed in 2026 remain incomplete. Heterogeneity in national legal frameworks, administrative practices, and applicant market strategies also introduced variation that could not be fully controlled. Finally, this study evaluates regulatory processes rather than downstream outcomes such as medicine availability, procurement timelines, or public health impact areas requiring additional investigation.

## 5. Conclusions

This study provides the first empirical evidence that a continent-wide scientific assessment and listing procedure for human medicinal products is feasible, efficient, and able to support reliance-based national regulatory decisions across Africa. Consolidated continental assessments were widely accepted by National Regulatory Authorities, although total timelines were shaped largely by administrative and governance sequencing rather than scientific review. Strengthening these procedural interfaces, together with embedding harmonised reliance pathways into national legislation and digital workflows, will be essential for achieving predictable, continent-wide access under the African Medicines Agency. Continued evaluation, including the impact on product availability and public health outcomes, will be necessary as the African Medicines Agency transitions from pilot to full operationalisation.

## Declarations

No declarations.

## Data Availability Statement

The data underlying this study contain regulatory and application-level information generated through the African Medicines Regulatory Harmonisation (AMRH) Programme and participating National Regulatory Authorities. Due to confidentiality obligations and data-sharing agreements, these data cannot be made publicly available. Aggregated and de-identified data supporting the findings of this study are provided within the manuscript and its Supplementary Information files. Access to underlying regulatory records may be considered upon reasonable request to AUDA-NEPAD, subject to applicable data-governance and confidentiality requirements.

## Acknowledgments

The authors acknowledge the contributions of the African Medicines Regulatory Harmonisation (AMRH) Steering Committee, Technical Committees including the Evaluation of Medicinal Products Technical Committee (EMP-TC), the Good Manufacturing Practices Technical Committee (GMP-TC), Continental Forum of Heads of Registration and Marketing Authorisation and the participating National Regulatory Authorities for their technical input and collaboration during the continental listing pilot. The authors also thank AUDA-NEPAD for institutional support and coordination of the pilot activities.

## Author Contributions

- **Conceptualization:** Alex Juma Ismail, Stuart Walker, Sam Salek
- **Methodology:** Alex Juma Ismail, Stuart Walker, Sam Salek
- **Formal Analysis:** Alex Juma Ismail
- **Data Curation:** Alex Juma Ismail
- **Writing – Original Draft:** Alex Juma Ismail
- **Writing – Review & Editing:** Alex Juma Ismail, Lerato Moeti, Delese Mimi Darko, Stuart Walker, Sam Salek
- **Supervision:** Delese Mimi Darko, Stuart Walker, Sam Salek

## Glossary of Terms

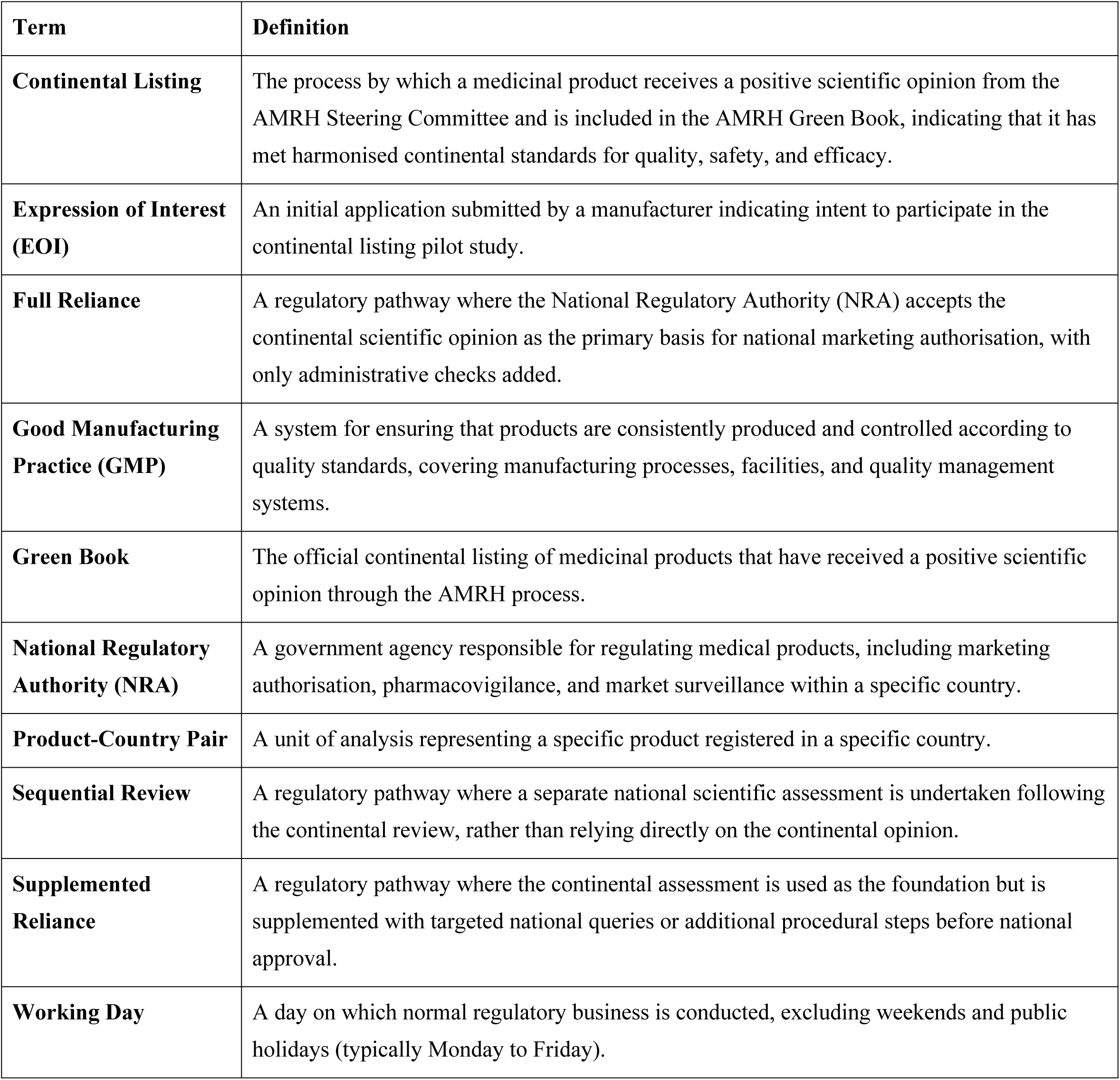

| Term | Definition |
| --- | --- |
| **Continental Listing** | The process by which a medicinal product receives a positive scientific opinion from the AMRH Steering Committee and is included in the AMRH Green Book, indicating that it has met harmonised continental standards for quality, safety, and efficacy. |
| **Expression of Interest (EOI)** | An initial application submitted by a manufacturer indicating intent to participate in the continental listing pilot study. |
| **Full Reliance** | A regulatory pathway where the National Regulatory Authority (NRA) accepts the continental scientific opinion as the primary basis for national marketing authorisation, with only administrative checks added. |
| **Good Manufacturing Practice (GMP)** | A system for ensuring that products are consistently produced and controlled according to quality standards, covering manufacturing processes, facilities, and quality management systems. |
| **Green Book** | The official continental listing of medicinal products that have received a positive scientific opinion through the AMRH process. |
| **National Regulatory Authority (NRA)** | A government agency responsible for regulating medical products, including marketing authorisation, pharmacovigilance, and market surveillance within a specific country. |
| **Product-Country Pair** | A unit of analysis representing a specific product registered in a specific country. |
| **Sequential Review** | A regulatory pathway where a separate national scientific assessment is undertaken following the continental review, rather than relying directly on the continental opinion. |
| **Supplemented Reliance** | A regulatory pathway where the continental assessment is used as the foundation but is supplemented with targeted national queries or additional procedural steps before national approval. |
| **Working Day** | A day on which normal regulatory business is conducted, excluding weekends and public holidays (typically Monday to Friday). |

## List of Abbreviations

AMA: African Medicines Agency
AMRH: African Medicines Regulatory Harmonisation
AU: African Union
AUDA-NEPAD: African Union Development Agency – New Partnership for Africa’s Development
CHMP: Committee for Medicinal Products for Human Use (EMA)
CRP: Collaborative Registration Procedure (WHO)
EAC: East African Community
ECOWAS: Economic Community of West African States
EMA: European Medicines Agency
EMP-TC: Evaluation of Medicinal Products Technical Committee
EOI: Expression of Interest
EU: European Union
GMP: Good Manufacturing Practice
GMP-TC: Good Manufacturing Practice Technical Committee
HPV: Human Papillomavirus
ICMRA: International Coalition of Medicines Regulatory Authorities
NRA: National Regulatory Authority
PQ: Prequalification (WHO)
SADC: Southern African Development Community
SAHPRA: South African Health Products Regulatory Authority
WAHO: West African Health Organization
WD: Working Day
WHO: World Health Organization

